# Victimization and witnessing of workplace bullying and physician-diagnosed physical and mental health and organizational outcomes: a cross-sectional study of a nationally representative sample in Japan

**DOI:** 10.1101/2022.03.10.22272191

**Authors:** Kanami Tsuno, Norito Kawakami, Akizumi Tsutsumi, Akihito Shimazu, Akiomi Inoue, Yuko Odagiri, Teruichi Shimomitsu

## Abstract

**Background:** Compared to the numerous reports on mental health outcomes of workplace bullying victims, research on organizational outcomes of witnesses and physical health outcomes of victims and witnesses is scarce. Therefore, the purpose of this study was to investigate the relationship between bullying victimization and witnessing and various physical and mental health outcomes and organizational outcomes such as sickness absence, work performance, and job satisfaction.

**Methods:** This study used cross-sectional data from a nationally representative, community-based sample of 5,000 Japanese residents aged 20-60. We analyzed data from 1,496 respondents after excluding those not working at the time of the survey and those with missing values. Workplace bullying, psychological distress, physical complaints, and job satisfaction were assessed with the New Brief Job Stress Questionnaire and work performance with the World Health Organization’s Health and Work Performance Questionnaire. In addition, subjective health status, physician-diagnosed mental or physical illness, and sickness absence were asked as one item. Hierarchical multiple regression analysis or Poisson regression analysis was conducted to assess the association between victimization/witnessing workplace bullying and health and organizational outcomes.

**Results:** Both victimization and witnessing workplace bullying were significantly associated with psychological distress, physical complaints, subjective poor health, physician-diagnosed mental disorders, and job dissatisfaction. Victimization of workplace bullying was further associated with physician-diagnosed respiratory diseases, sickness absence (≥7 days), and poor work performance. Victims were absent from work for 4.5 more sick days and had 11.2% lower work performance than non-victims.

**Conclusions:** The results showed that both victimization and witnessing workplace bullying were significantly associated with physical and mental outcomes and various organizational outcomes. Organizations should implement further measures to prevent personal and organizational losses due to workplace bullying.

## Introduction

Workplace bullying is one of the most severe psychosocial stressors at work. Several meta-analyses and systematic reviews have already been conducted to confirm the association between exposure to workplace bullying and various mental health outcomes, such as depression or anxiety (1, 2), post-traumatic stress disorder (PTSD) (3), sleep (4), and suicidal ideation (5). These studies clearly show that workplace bullying has unquestionably harmed the mental health of the victims.

Compared to the numerous reports on mental health outcomes of workplace bullying, few studies have focused on physical health outcomes (6). Disease-level physical health outcomes of workplace bullying have been reported, including cardiovascular disease (7, 8), type 2 diabetes (9), and fibromyalgia (10). By contrast, an association between workplace bullying and other chronic diseases, such as respiratory and gastrointestinal diseases, has not been thoroughly investigated to the best of our knowledge. For instance, Kivimaki, Elovainio (11) reported that a higher proportion of victims had chronic diseases among Finnish hospital employees (n = 5,655). However, they did not report which chronic diseases they had more than non-victims. To date, only one cross-sectional study has reported that bullying was a predictor of asthma, a respiratory disease, in the Peruvian sample of cleaners (n = 199) (12). Although a recent study reported that workplace bullying was associated with increased doctor visits, the diagnosis is unknown (13). On the other hand, several qualitative studies have reported that victims of workplace bullying had symptoms of asthma or gastric ulcers (14, 15). Therefore, more quantitative study is needed to investigate the association between workplace bullying and various physical diseases, including respiratory diseases or digestive diseases.

The primary organizational outcomes of workplace bullying are absenteeism, turnover, and work performance. Meta-analytic studies have found significant associations between workplace bullying and sickness absence and poor work performance (16, 17). However, most of the studies that have examined the work performance of bullying victims have only calculated the correlation coefficient between bullying and work performance without using standardized measures (17, 18). For example, although Kivimaki, Elovainio (11) reported that victims had a 26% higher risk of taking sickness absence, they did not report how many more days the victims took off for sickness absence than non-victims. To calculate workplace bullying costs (19, 20), clarifying the difference between victims and non-victims is essential. Thus, this study investigates the relationship between bullying and other organizational outcomes, such as job satisfaction, and examines how many more days victims take off as sickness absence and how many percent less they work than non-victims.

Few studies have examined witnesses’ health and organizational outcomes after adjusting for exposure to bullying. Most studies have included some victims among the witnesses, contributing to overestimating the health effects of witnessing workplace bullying (21). Therefore, when investigating witness health outcomes, researchers have to exclude bullied people from witnesses or control for the experience of being bullied to see a “pure” effect of witnessing bullying. Although a multilevel study has reported that department-level bullying can affect subsequent psychological distress and intention to leave, even when controlling for individual exposure to bullying (22), no studies have examined the association between witnessing bullying and organizational outcomes such as sickness absence and work performance, to the best of our knowledge.

To sum up, previous research has focused primarily on mental health outcomes of bullying victimization. Furthermore, most bullying studies used specific workers, such as health care workers, and cannot be generalized to the general working population. Therefore, the purpose of this study was to investigate the association between bullying victimization and witnessing and various physical and mental health outcomes, such as physician-diagnosed physical and mental disorders, subjective health, and physical complaints, as well as organizational outcomes, such as sickness absence, work performance, and job satisfaction, using in a nationally representative sample in Japan.

## Methods

### Participants

This cross-sectional study was conducted in 2010 for a nationally representative community-based sample of 5,000 Japanese residents between the ages of 20 and 60. The details of the random sampling were described elsewhere (23). A total of 2,384 agreed to participate and completed the questionnaire (response rate: 47.7%). After excluding 751 respondents who were not working at the time of the survey and 137 respondents who had missing responses on sex, age, education, occupation, employment, workplace bullying, subjective health status, sickness absence, job satisfaction, the data from 1,496 respondents were analyzed in this study.

### Ethics Statement

The Ethical Committee of the Graduate School of Medicine/Faculty of Medicine, The University of Tokyo, reviewed and approved this study’s aims and procedures before conducting the survey (#2953).

### Measures

#### Workplace bullying

Workplace bullying was assessed using a self-labeling method without a definition, using the New Brief Job Stress Questionnaire (New BJSQ) (24, 25). First, respondents were asked whether they experienced bullying at the survey time. The respondents who answered “1 = very much so” or “2 = moderately so” were defined as “victims” (23). Respondents were also asked whether there are people who are bullied or harassed in their workplace, and those who answered “1 = very much so” or “2 = moderately so” were defined as “witnesses.” Three categories were created from these two questions: “not bullied nor witnessed,” “not bullied but witnessed,” and “bullied” since both experiencing and witnessing bullying have been reported as risk factors for adverse health outcomes (22).

#### Health outcomes

Five aspects measured psychological distress: vigor (three items), anger-irritability (three items), fatigue (three items), anxiety (three items), depression (six items) using an 18-item scale of the New BJSQ (24). Each item sample is “I have been full of energy (reverse item)” (vigor), “I have felt angry” (anger-irritability), “I have felt extremely tired” (fatigue), “I have felt worried or insecure” (anxiety), and “I have felt sad” (depression). A four-point Likert-style response option was used: “almost never = 1” to “almost always= 4.” Average scores of 18 items were calculated for analysis. Higher scores mean having greater psychological distress.

Physician-diagnosed mental disorders were measured by asking whether the individual has received treatment for a mental disorder, including depression. Those who answered “yes” were determined to have a mental disorder. In Japan, “treatment” refers to a medical treatment based on a physician’s diagnosis and is performed only by the physician. The Medical Practitioners Law strictly prohibits other medical personnel from performing medical treatment, including medication prescription. Therefore, in this study, “physician-diagnosed mental disorders” refer to mental disorders that a physician is currently treating.

For physician-diagnosed physical diseases, respondents were asked, “Are you currently receiving treatment for any of the following diseases or symptoms?” and answered “yes” or “no” to chronic diseases such as cardiovascular diseases (i.e., hypertension, heart disease, stroke), diabetes, respiratory diseases (i.e., asthma, chronic bronchitis), digestive diseases (i.e., stomach ulcer, liver disease), and orthopedic diseases (i.e., back pain). In this survey, physician-diagnosed physical diseases refer to diseases currently being treated by a physician.

Physical complaints were measured by an 11-item of the New BJSQ (24). The item samples are “I have experienced headaches” and “I have felt dizzy.” Response options were the same as for the psychological distress scale of the BJSQ. The higher the score, the greater the physical complaints.

Subjective health status was measured with a single item, “Overall, how was your health during the past month?” Response options ranged from “not good at all = 1” to “perfect = 6” and those who answered “perfect,” “very good,” or “good” classified as “good,” and those who answered “not so good,” “not good,” or “not good at all” classified as “poor.”

#### Organizational outcomes

To measure sickness absence, we asked, “In the past year, how many days in total did you take off from work due to health problems?” Two categories were created from this question: sickness absence (≥1 day) and sickness absence (≥7 days).

A single item measured work performance (relative presenteeism) from the World Health Organization’s Health and Work Performance Questionnaire (WHO-HPQ) (25, 26). The respondents were asked, “On a scale from 0 to 10 where 0 is the worst work performance anyone could have at your job and 10 is the performance of a top worker, how would you rate your overall work performance on the days you worked during the past four weeks (28 days)?” Again, response options were 0 to 10, and a higher score means having more excellent work performance.

Job satisfaction was measured by one item of the New BJSQ (24). Response options ranged from “dissatisfied = 1” to “satisfied = 4,” with those who answered “satisfied” or “somewhat satisfied” classified as “satisfied” and those who answered “somewhat dissatisfied” or “dissatisfied” classified as “dissatisfied.”

#### Other covariates

As individual and socioeconomic status (SES) characteristics, sex, age, education, household income during the past year, occupation, and employment were asked to the respondents. Then, dummy variables were created for analyses: sex (male = 1, female = 0), age (under 29 =1, over 30 =0), education (high school graduates or below= 1, college graduates or above =0), household income (less than 2.5 million yen [equivalent to < US$22,000, if 1$ = ¥115] = 1, over 250 million yen = 0), occupation (manager =1, others =0), and employment (permanent =1, others =0).

### Statistical Analysis

First, Spearman’s correlation coefficients were calculated between all variables. Second, mean values of continuous variables including psychological distress, physical complaints, sickness absence, and work performance were compared among victims, witnesses, and non-victims/non-witnesses by analysis of variance (ANOVA). Then, hierarchical multiple regression analyses were conducted to examine the relationship between experienced or witnessed bullying at work and psychological distress, physical complaints, and work performance. Finally, we conducted Poisson regression analyses to examine the relationship between workplace bullying and categorical health outcomes, including physician-diagnosed diseases and subjective health and organizational outcomes, including sickness absence (≥1 or ≥7) and job satisfaction. Prevalence ratios (PRs) and 95% Confidence Intervals (CIs) were calculated, adjusting for individual characteristics (sex and age) and SES variables (education, household income, occupation, and employment status). The 2-tailed *p-*value for statistical significance to see the differences among each social indicator was set at 0.05. All analyses were conducted using SPSS 27.0 for Windows.

## Results

### Characteristics of the Respondents

Table 1 shows the characteristics of the respondents of this study. Most of the respondents were males, 40-49 years old, graduated high school or below, had a household income between ¥2.50 million and ¥4.99 million, had professional or technical jobs, and were permanent (full-time) employees. Six percent of the respondent had experienced workplace bullying, and ten percent had not been bullied but witnessed bullying at the workplace. Approximately 60% of the respondents rated their health as “good,” had at least one day of sickness absence during the past year, and rated their job satisfaction as “satisfied.”

**Table 1.** Characteristics of respondents in this study (N = 1,496).

|  | n | % |  | n | % |
| --- | --- | --- | --- | --- | --- |
| **Individual and socioeconomic characteristics** |  |  | **Health outcomes** |  |  |
| Sex |  |  | Physician-diagnosed mental disorders |  |  |
| Male | 781 | 52.2 | Yes | 32 | 2.1 |
| Female | 715 | 47.8 | No | 1464 | 97.9 |
| Age |  |  | Physician-diagnosed cardiovascular diseases |  |  |
| < 30 | 234 | 15.6 | Yes | 143 | 9.6 |
| 30-39 | 422 | 28.2 | No | 1353 | 90.4 |
| 40-49 | 428 | 28.6 | Physician-diagnosed diabetes |  |  |
| ≥ 50 | 412 | 27.5 | Yes | 63 | 4.2 |
| Education |  |  | No | 1433 | 95.8 |
| High school graduate or below | 679 | 45.4 | Physician-diagnosed respiratory diseases |  |  |
| Vocational school/college graduate | 401 | 26.8 | Yes | 36 | 2.4 |
| University/graduate school graduate | 416 | 27.8 | No | 1460 | 97.6 |
| Household income (million yen) |  |  | Physician-diagnosed digestive diseases |  |  |
| < 2.50 | 125 | 8.4 | Yes | 98 | 6.6 |
| 2.50-4.99 | 453 | 30.3 | No | 1398 | 93.4 |
| 5.00-7.49 | 395 | 26.4 | Physician-diagnosed orthopedic diseases |  |  |
| 7.50-9.99 | 240 | 16.0 | Yes | 191 | 12.8 |
| ≥ 10.00 | 152 | 10.2 | No | 1305 | 87.2 |
| Unknown | 131 | 8.8 | Physician-diagnosed other chronic diseases |  |  |
| Occupation |  |  | Yes | 201 | 13.4 |
| Managers | 144 | 9.6 | No | 1295 | 86.6 |
| Professionals or technicians | 338 | 22.6 | Subjective health status |  |  |
| Clerks | 281 | 18.8 | Good | 929 | 62.1 |
| Sales workers | 160 | 10.7 | Poor | 567 | 37.9 |
| Service workers | 151 | 10.1 |  |  |  |
| Production workers and laborers | 225 | 15.0 | **Organizational outcomes** |  |  |
| Others | 197 | 13.2 | Sickness absence (≥1) |  |  |
| Employment contract |  |  | Yes | 895 | 59.8 |
| Permanent | 969 | 64.8 | No | 601 | 40.2 |
| Temporary/contract/part-time | 477 | 31.9 | Sickness absence (≥7) |  |  |
| Others | 50 | 3.3 | Yes | 417 | 27.9 |
| **Workplace bullying** |  |  | No | 1079 | 72.1 |
| Not bullied nor witnessed | 1254 | 83.8 | Job satisfaction |  |  |
| Not bullied but witnessed | 151 | 10.1 | Satisfied | 918 | 61.4 |
| Bullied | 91 | 6.1 | Dissatisfied | 578 | 38.6 |

### Correlations between Variables

Table 2 shows Spearman’s correlation coefficients between all variables in this study. Experiencing workplace bullying was significantly and positively associated with younger age, low household income, psychological distress, physician-diagnosed mental disorders, physician-diagnosed respiratory diseases, physical complaints, subjective poor health, sickness absence, and job dissatisfaction, while significantly and negatively associated with work performance. Witnessing bullying at the workplace was also significantly and positively associated with psychological distress, physician-diagnosed mental disorders, physical complaints, subjective poor health, sickness absence, and job dissatisfaction.

**Table 2.** Spearman’s correlations between variables.

| Variables † | 1 | 2 | 3 | 4 | 5 | 6 | 7 | 8 | 9 | 10 | 11 | 12 | 13 | 14 | 15 | 16 | 17 | 18 | 19 | 20 |
| --- | --- | --- | --- | --- | --- | --- | --- | --- | --- | --- | --- | --- | --- | --- | --- | --- | --- | --- | --- | --- |
| 1 Age (under 29 = 1) | 1 |  |  |  |  |  |  |  |  |  |  |  |  |  |  |  |  |  |  |  |
| 2 Sex (male = 1) | -.07** | 1 |  |  |  |  |  |  |  |  |  |  |  |  |  |  |  |  |  |  |
| 3 Education (high school = 1) | -.10** | -.03 | 1 |  |  |  |  |  |  |  |  |  |  |  |  |  |  |  |  |  |
| 4 Household income (< 250 = 1) | .04 | -.09** | .06* | 1 |  |  |  |  |  |  |  |  |  |  |  |  |  |  |  |  |
| 5 Occupation (manager = 1) | -.13** | .25** | -.04 | -.07** | 1 |  |  |  |  |  |  |  |  |  |  |  |  |  |  |  |
| 6 Employment (permanent = 1) | .07** | .46** | -.13** | -.18** | .12** | 1 |  |  |  |  |  |  |  |  |  |  |  |  |  |  |
| 7 Psychological distress | .08** | .08** | -.03 | .05* | -.04 | .13** | 1 |  |  |  |  |  |  |  |  |  |  |  |  |  |
| 8 Mental disorders (yes = 1) | -.03 | .04 | .02 | .06* | .03 | .04 | .08** | 1 |  |  |  |  |  |  |  |  |  |  |  |  |
| 9 Cardiovascular diseases (yes = 1) | -.13** | .08** | .07* | .03 | .14** | -.00 | -.02 | -.00 | 1 |  |  |  |  |  |  |  |  |  |  |  |
| 10 Diabetes (yes = 1) | -.06* | .11** | .00 | -.02 | .08** | .02 | -.01 | .04 | .22** | 1 |  |  |  |  |  |  |  |  |  |  |
| 11 Respiratory diseases (yes = 1) | -.01 | .00 | -.01 | .00 | -.01 | -.02 | .05 | .01 | .04 | -.01 | 1 |  |  |  |  |  |  |  |  |  |
| 12 Digestive diseases (yes = 1) | -.07** | .02 | .03 | -.01 | .02 | -.03 | .02 | .05* | .10** | .08** | .08** | 1 |  |  |  |  |  |  |  |  |
| 13 Orthopedic diseases (yes = 1) | -.04 | -.02 | .06* | .00 | .01 | -.00 | .08** | -.04 | .05 | .06* | .02 | .03 | 1 |  |  |  |  |  |  |  |
| 14 Other chronic diseases (yes = 1) | -.03 | -.07* | -.02 | .00 | -.04 | -.05* | .06* | .01 | -.04 | -.02 | .00 | -.03 | .03 | 1 |  |  |  |  |  |  |
| 15 Physical complaints | .03 | -.08** | .03 | .06* | -.03 | .02 | .61** | .07** | .01 | .05* | .08** | .15** | .17** | .12** | 1 |  |  |  |  |  |
| 16 Poor subjective health (poor = 1) | .04 | -.03 | .03 | .06* | -.03 | .02 | .44** | .14** | .08** | .10** | .08** | .14** | .16** | .15** | .46** | 1 |  |  |  |  |
| 17 Sickness absence | .03 | -.02 | .02 | .02 | -.04 | .02 | .10** | .08** | .05* | .04 | .04 | .07** | .12** | .09** | .13** | .13** | 1 |  |  |  |
| 18 Work performance | -.18** | .02 | .07** | -.02 | .01 | -.08** | -.21** | -.04 | .03 | .02 | -.02 | .03 | .03 | -.04 | -.08** | -.10** | -.07** | 1 |  |  |
| 19 Job dissatisfaction (dissatisfied = 1) | .10** | .05* | .01 | .01 | -.05* | .09** | .49** | .03 | -.02 | -.01 | .06* | .03 | .06* | .07** | .28** | .26** | .08** | -.20** | 1 |  |
| 20 Bullied at work (yes = 1) | .06* | .00 | -.03 | .10** | -.03 | -.01 | .21** | .08** | -.02 | .02 | .07** | .00 | .03 | .04 | .16** | .16** | .07** | -.08** | .17** | 1 |
| 21 Witnessed bullying at work (yes = 1) | -.03 | .01 | -.03 | .01 | -.05 | -.00 | .18** | .06* | -.01 | -.04 | .02 | .03 | -.00 | .04 | .10** | .10** | .05 | -.01 | .12** | -.09** |

SD: standard deviation.
\* $p < .05$ ; \*\* $p < .01$ .

### Comparison of the Mean Scores of Psychological Distress, Physical Complaints, Sickness Absence, and Work Performance

Table 3 shows the comparison of the mean values of continuous outcome variables among victims (n=91), witnesses (n=151), and non-bullied/non-witnessed respondents (n=1,254) by ANOVA. The highest scores in victims and second-highest scores in witnesses were observed in psychological distress and physical complaints. Victims reported significantly lower work performance than non-bullied/non-witnessed respondents; the difference of the scores was 0.75, which means an 11.2% difference between victims and non-victims (0.75/6.72*100). Witnesses also reported significantly lower work performance than non-bullied/witnessed respondents. In contrast, the mean days of sickness absence were not significantly different among victims, witnessed, and non-bullied/non-witnessed respondents, although the difference was 4.5 days between victims and non-victims.

**Table 3.** Mean values of psychological distress, physical complaints, sickness absence, and work performance of bullied respondents (n = 91) and witnesses (n = 151) compared with non-bullied/witnessed respondents (n = 1,254): ANOVA.

| **Variables:** | Mean | SD | *p* value |
| --- | --- | --- | --- |
| Psychological distress |  |  | < 0.001 |
| Not bullied nor witnessed | 2.07 *ab | 0.57 |  |
| Not bullied but witnessed | 2.47 *ac | 0.56 |  |
| Bullied | 2.73 *bc | 0.68 |  |
| Physical complaints |  |  | < 0.001 |
| Not bullied nor witnessed | 1.73 *ab | 0.51 |  |
| Not bullied but witnessed | 1.91 *ac | 0.52 |  |
| Bullied | 2.16 *bc | 0.64 |  |
| Sickness absence (days) |  |  | 0.230 |
| Not bullied nor witnessed | 8.42 | 23.87 |  |
| Not bullied but witnessed | 8.84 | 21.63 |  |
| Bullied | 12.93 | 31.98 |  |
| Work performance |  |  | < 0.001 |
| Not bullied nor witnessed | 6.72 *a | 1.70 |  |
| Not bullied but witnessed | 6.61 *b | 1.77 |  |
| Bullied | 5.97 *ab | 2.23 |  |

\* $p < .05$ . by Bonferroni.

### Relationship between Workplace Bullying and Psychological Distress, Physical Complaints, and Work Performance

Table 4 shows the results of hierarchical regression analyses of bullying and continuous outcome variables. In Step 2 where sex, age, education, household income, occupation, and employment were entered, both experiencing and witnessing workplace bullying were significantly and positively associated with psychological distress (b = 0.64; 0.40, *p* < 0.001), physical complaints (b = 0.43; 0.19, *p* < 0.001); significantly and negatively associated with work performance (b = -0.68; -0.14, *p* < 0.001). However, the regression coefficients were larger in the association between bullying victimization and outcomes than witness and outcomes.

**Table 4.**
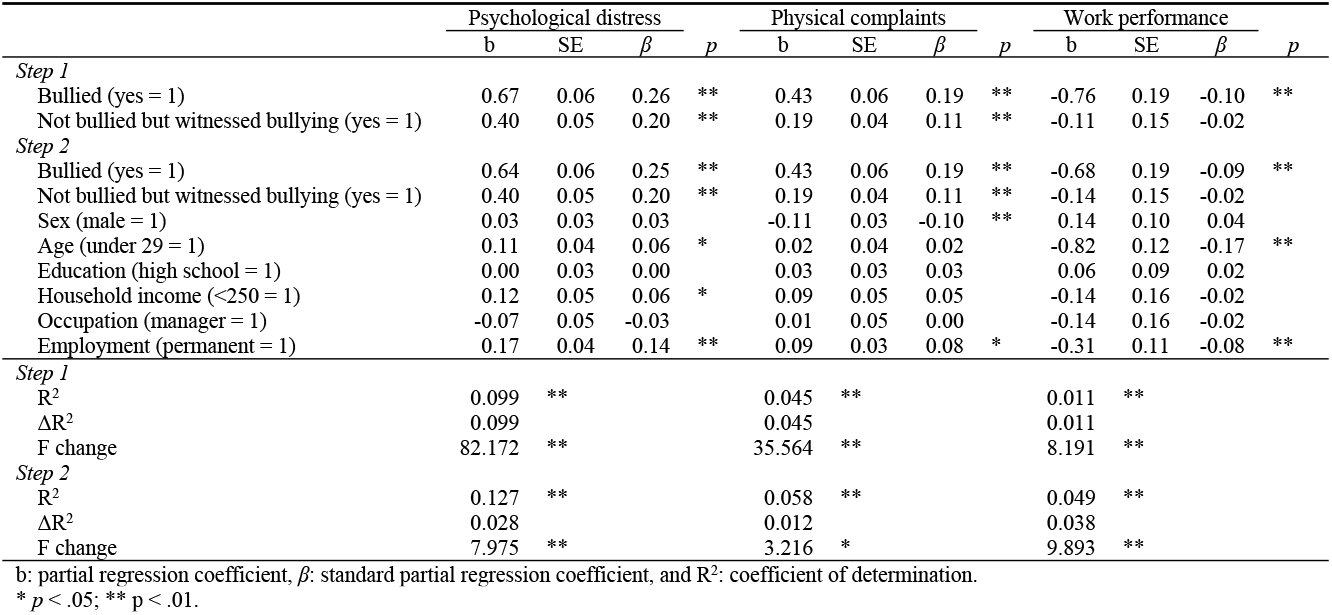
Hierarchical multiple regression of bullying and psychological distress, physical complaints, and work performance.

|  | Psychological distress |  |  |  | Physical complaints |  |  |  | Work performance |  |  |  |
| --- | --- | --- | --- | --- | --- | --- | --- | --- | --- | --- | --- | --- |
| | b | SE | $\beta$ | p | b | SE | $\beta$ | p | b | SE | $\beta$ | p |
| *Step 1* |  |  |  |  |  |  |  |  |  |  |  |  |
| Bullied (yes = 1) | 0.67 | 0.06 | 0.26 | ** | 0.43 | 0.06 | 0.19 | ** | -0.76 | 0.19 | -0.10 | ** |
| Not bullied but witnessed bullying (yes = 1) | 0.40 | 0.05 | 0.20 | ** | 0.19 | 0.04 | 0.11 | ** | -0.11 | 0.15 | -0.02 |  |
| *Step 2* |  |  |  |  |  |  |  |  |  |  |  |  |
| Bullied (yes = 1) | 0.64 | 0.06 | 0.25 | ** | 0.43 | 0.06 | 0.19 | ** | -0.68 | 0.19 | -0.09 | ** |
| Not bullied but witnessed bullying (yes = 1) | 0.40 | 0.05 | 0.20 | ** | 0.19 | 0.04 | 0.11 | ** | -0.14 | 0.15 | -0.02 |  |
| Sex (male = 1) | 0.03 | 0.03 | 0.03 |  | -0.11 | 0.03 | -0.10 | ** | 0.14 | 0.10 | 0.04 |  |
| Age (under 29 = 1) | 0.11 | 0.04 | 0.06 | * | 0.02 | 0.04 | 0.02 |  | -0.82 | 0.12 | -0.17 | ** |
| Education (high school = 1) | 0.00 | 0.03 | 0.00 |  | 0.03 | 0.03 | 0.03 |  | 0.06 | 0.09 | 0.02 |  |
| Household income (<250 = 1) | 0.12 | 0.05 | 0.06 | * | 0.09 | 0.05 | 0.05 |  | -0.14 | 0.16 | -0.02 |  |
| Occupation (manager = 1) | -0.07 | 0.05 | -0.03 |  | 0.01 | 0.05 | 0.00 |  | -0.14 | 0.16 | -0.02 |  |
| Employment (permanent = 1) | 0.17 | 0.04 | 0.14 | ** | 0.09 | 0.03 | 0.08 | * | -0.31 | 0.11 | -0.08 | ** |
| *Step 1* |  |  |  |  |  |  |  |  |  |  |  |  |
| R<sup>2</sup> | 0.099 | ** |  |  | 0.045 | ** |  |  | 0.011 | ** |  |  |
| $\Delta R^2$ | 0.099 | | | | 0.045 | | | | 0.011 | | | |
| F change | 82.172 | ** |  |  | 35.564 | ** |  |  | 8.191 | ** |  |  |
| *Step 2* |  |  |  |  |  |  |  |  |  |  |  |  |
| R<sup>2</sup> | 0.127 | ** |  |  | 0.058 | ** |  |  | 0.049 | ** |  |  |
| $\Delta R^2$ | 0.028 | | | | 0.012 | | | | 0.038 | | | |
| F change | 7.975 | ** |  |  | 3.216 | * |  |  | 9.893 | ** |  |  |

b: partial regression coefficient, $\beta$ : standard partial regression coefficient, and R<sup>2</sup>: coefficient of determination.
\* $p < .05$ ; \*\* $p < .01$ .

### Relationship between Workplace Bullying and physician-diagnosed Psychical and Mental Disorders under Treatment, Subjective Health, Sickness Absence, and Job Satisfaction

Table 5 shows the results of Poisson regressions of bullying and categorical health and organizational outcome variables. Both an exposure to workplace bullying and witnessing bullying at the workplace were significantly associated with subjective poor health (PR: 2.00 [95%CI: 1.53 to 2.61]; 1.52 [1.19 to 1.94]), physician-diagnosed mental disorders (PR: 3.93 [1.55 to 10.00)]; 2.91 [1.22 to 6.92)]), and job dissatisfaction (PR: 1.99 [1.53 to 2.60]; 1.61 [1.27 to 2.04]), after adjusting for individual characteristics and SES variables. In addition, exposure to workplace bullying was significantly associated with sickness absence (≥ 7) (PR: 1.56 [1.10 to 2.19)]) and physician-diagnosed respiratory 1 diseases (PR: 3.33 [1.35 to 8.23]) in the adjusted model.

**Table 5.** Workplace bullying and physician-diagnosed physical and mental disorders, subjective health, sickness absence, and job satisfaction: Poisson regression analysis.

| Outcome variables: | Crude |  | Adjusted † |  |
| --- | --- | --- | --- | --- |
|  | Not bullied but witnessed | Bullied | Not bullied but witnessed | Bullied |
|  | PRs (95% CI) | PRs (95% CI) | PRs (95% CI) | PRs (95% CI) |
| Mental disorders | **2.91 (1.22 to 6.92)** | **3.93 (1.55 to 10.00)** | **2.91 (1.22 to 6.92)** | **3.93 (1.55 to 10.00)** |
| Cardiovascular diseases | 0.87 (0.49 to 1.54) | 0.78 (0.37 to 1.67) | 0.90 (0.51 to 1.60) | 0.84 (0.39 to 1.82) |
| Diabetes | 0.45 (0.14 to 1.45) | 1.26 (0.50 to 3.14) | 0.47 (0.15 to 1.49) | 1.33 (0.53 to 3.34) |
| Respiratory diseases | **2.90 (1.23 to 6.86)** | **4.15 (1.67 to 10.34)** | 1.62 (0.62 to 4.24) | **3.33 (1.35 to 8.23)** |
| Digestive diseases | 1.36 (0.76 to 2.45) | 1.05 (0.46 to 2.41) | 1.36 (0.75 to 2.44) | 1.14 (0.50 to 2.64) |
| Orthopedic diseases | 1.00 (0.62 to 1.60) | 1.31 (0.77 to 2.23) | 1.01 (0.63 to 1.62) | 1.39 (0.82 to 2.37) |
| Other chronic diseases | 1.36 (0.91 to 2.07) | 1.49 (0.90 to 2.46) | 1.33 (0.87 to 2.01) | 1.51 (0.91 to 2.51) |
| Poor subjective health | **1.51 (1.19 to 1.93)** | **2.04 (1.56 to 2.66)** | **1.52 (1.19 to 1.94)** | **2.00 (1.53 to 2.61)** |
| Sickness absence ($\geq 1$) | 1.11 (0.90 to 1.36) | 1.21 (0.93 to 1.56) | 1.11 (0.90 to 1.37) | 1.19 (0.92 to 1.54) |
| Sickness absence ($\geq 7$) | 1.14 (0.84 to 1.55) | **1.53 (1.09 to 2.15)** | 1.14 (0.83 to 1.55) | **1.56 (1.10 to 2.19)** |
| Job dissatisfaction | **1.60 (1.27 to 2.03)** | **2.04 (1.57 to 2.65)** | **1.61 (1.27 to 2.04)** | **1.99 (1.53 to 2.60)** |

† Individual characteristics (sex and age) and SES (education, household income, occupation, and employment status) adjusted in the model.
Reference group: not exposed nor witnessed workplace bullying.
Bold figures refer to significant results.

## Discussion

The current study aimed to investigate the association between experiencing and witnessing bullying at work and various health and organizational outcomes in a nationally representative sample in Japan. The study results revealed victimization of workplace bullying was significantly associated with psychological distress, physician-diagnosed mental disorders, physician-diagnosed respiratory diseases, physical complaints, subjective poor health, sickness absence (≥ 7), lower work performance, and job dissatisfaction, after adjusting for potential confounders. Witnessing bullying was also associated with psychological distress, physician-diagnosed mental disorders, physical complaints, subjective poor health, and job dissatisfaction. In addition, victims had 4.5 more days of sickness absence than non-victims, although it was not statistically significant. In contrast, victims had 11.2% significantly lower work performance than non-victims. Overall, our study results suggest that experiencing and witnessing bullying is associated with various health and organizational outcomes. In addition, this study added to the literature that bullying experience was associated with physician-diagnosed diseases, including mental disorders and respiratory diseases.

Workplace bullying was associated with having physician-diagnosed mental disorders under treatment, in addition to the association with psychological distress and physical complaints that were measured by a scale. Additionally, witnessing bullying was also associated with physician-diagnosed mental disorders under treatment. Although a meta-analysis study reported workplace bullying was related to depressive symptoms, anxiety symptoms, PTSD symptoms, and psychological complaints, few studies have focused on physician-diagnosed mental disorders (2, 7). People who sought psychiatric treatments could have more deterioration in their social functioning than people with non-clinical psychological distress. Thus, physician-diagnosed mental disorders may be a more relevant outcome to assess the health and social impact of workplace bullying. Thus, although causality cannot be determined since having mental disorders was also reported as a predictor of workplace bullying (2, 27), our study results added the literature that exposure to workplace bullying is associated with clinical mental illness in a representative working sample in Japan.

Our finding that exposure to workplace bullying was significantly associated with physician-diagnosed respiratory diseases under treatment was relatively “new” to this field. However, this coincides with an empirical study that reported the association between workplace bullying and asthma among Peruvian cleaners (12) or a qualitative study that reported victims had symptoms of asthma (14). This is not surprising because stress triggers clinically significant bronchoconstriction or exacerbation of asthma (28, 29). Moreover, since long-term exposure to stress (life events and appraisals of threat and manageability) can increase susceptibility to respiratory diseases (30), workplace bullying may also trigger or exacerbate such illnesses.

The study results show that both victimization and witness to workplace bullying were associated with subjective poor health and job dissatisfaction. This is in line with the studies that reported exposure to workplace bullying influences job satisfaction in Belgian, Norwegian, Italian, and Spanish samples (31-33). Although little study investigated the effect of witnessing workplace bullying on individual and organizational outcomes (34, 35), a recent study confirmed the adverse effects of witnessing bullying on job satisfaction, organizational commitments, and turnover intentions after controlling for witnesses’ own experiences of being bullied (36). Our study results also confirmed this association after excluding those who were bullied from witnesses, indicating the existence of workplace bullying influences witnesses’ motivation and organizational commitment. As previously reported in the longitudinal study, the existence of bullying at the department level increases employees’ subsequent psychological distress and intention to leave (22). Our study also showed that witnesses (those who were not bullied but witnessed) had higher psychological distress and physical complaints scores than those not bullied nor witnessed, while the highest scores were observed among those who were bullied. In contrast, no significant difference was found in sickness absence and work performance between those who did not experience or witness bullying. Thus, further studies are needed to clarify this association.

The study found that victims had 4.5 more days of absenteeism and 11.2% lower work performance in the previous year than non-victims, consistent with studies that reported an association between exposure to bullying and absenteeism and work performance. (11, 16, 17). Interestingly, this difference in productivity or sickness absence was comparable to a nationwide survey in the UK (20). They reported that bullying victims were 7% less productive and 7 days more off work during the previous year than employees who were neither bullied nor witnessed (20). This indicates that workplace bullying affects the productivity of the organization itself and increases organizational costs to replace those who are on sick leave. To prevent individual and organizational losses due to workplace bullying, organizations need to implement further anti-bullying measures.

Several limitations need to be noted. First, the nature of the cross-sectional design precludes determining causality. As reported in several studies, mental health status also predicts bullying victimization (2, 27). This nature of the association between workplace bullying and mental health may have contributed to the overestimation of the association between bullying and mental disorders in the current cross-sectional study. It is unclear whether physical health status also predicts workplace bullying victimization, but this possibility cannot be ruled out. Longitudinal studies are needed to clarify workplace bullying and various health outcomes and organizational outcomes. Second, this study did not ask for the name of the diagnosis for which the respondent was receiving treatment. Since the disease severity varies, future research should focus on the name of the diagnosis and the severity of the disease. Third, we used a self-labeling method to measure workplace bullying, which has been previously reported to underestimate the prevalence of workplace bullying (37). Fourth, the possible measurement error may have contributed to underestimating (or overestimating) the association between bullying and sickness absence since sickness absence days were obtained by self-report in this study. If possible, the use of the organizations’ official sick leave data would allow for a more objective investigation of the victims’ sick leave.

Despite some limitations, the strength of this study is the use of a representative Japanese sample, and the results of this study can be generalized to the general Japanese workforce population. Another strength is that we are investigating various clinical levels of physician-diagnosed diseases. As mentioned in the introduction, quantitative studies on workplace bullying and physical diseases are still scarce (6). The author believes that this study will encourage future research in this field, as it showed a link between workplace bullying and physician-diagnosed diseases such as mental disorders and respiratory diseases. Finally, another strength of this study is that it focuses on both victims and witnesses of workplace bullying. As previous studies have suggested, witnesses of bullying also suffer from mental illness (22), but this is often neglected in research. Future research should focus on the various health problems of both victims and witnesses of bullying in order to understand the adverse effects of workplace bullying as a whole.

## Conclusions

The study found that victimization and witnessing workplace bullying were significantly associated with psychological distress, physician-diagnosed mental disorders, physical complaints, subjective poor health, and job dissatisfaction. Furthermore, workplace bullying victimization was associated with physician-diagnosed respiratory disorders, sickness absence (≥7), and poor work performance. To prevent individual and organizational losses due to workplace bullying, organizations need to implement further anti-bullying measures.

## Data Availability

The data underlying the results presented in the study are available from the first author (Kanami Tsuno).

## Acknowledgments

The authors thank Dr. Toru Yoshikawa (Department of Research, The Institute for Science of Labour, Kawasaki, Japan) for his support in conducting this study.

## Notes

### Competing Interest Statement

The authors have declared no competing interest.

### Funding Statement

This study was supported by the Ministry of Health, Labour and Welfare, Japan (#H21-Rodo-Ippan-001) and the Japan Society for the Promotion of Science (#19K19439). The funders had no role in study design, data collection and analysis, decision to publish, or preparation of the manuscript.

